# Pre-Deployment Cardiovascular Risk Assessment and Outcomes among United States Antarctic Program Participants: Insights from Contemporary Risk Models

**DOI:** 10.64898/2026.10.14.26365580

**Authors:** Daniel LeBlanc, Abdul Qadeer, Shiva Raju Gollapally Krishna, Andrew Forrester, Anjni Patel, Daniel Shi, Eric Nagueh, Jeremy Hsu, Neel Bhobe, Diann Gaalema, James McKeith, Hani Jneid, Joseph Allencherril

**Affiliations:** University of Texas Medical Branch, 301 University Boulevard, Galveston, TX 77555, USA; Center for Polar Medical Operations, 301 University Boulevard, Galveston, TX 77555, USA

**Keywords:** Cardiovascular Risk Assessment, Risk Stratification, Antarctic Regions, PREVENT Equations, Occupational Health

## Abstract

**Background:** Cardiovascular (CV) screening is critical for personnel deploying to Antarctica, where geographic isolation and limited medical care make the management of acute cardiovascular events challenging. The utility of contemporary risk prediction tools such as the AHA PREVENT equations in this unique population remains uncertain.

**Methods:** We retrospectively analyzed 154 individuals referred for enhanced CV evaluation before deployment to the U.S. Antarctic Program (USAP). Participants were evaluated across three temporal cohorts (2018-2020, 2021-2023, and 2024-2026). Demographics, CV risk factors, cardiac testing, and deployment clearance outcomes were collected. Risk was estimated using the Framingham Risk Score (FRS), AHA/ACC Pooled Cohort Equations (PCE), and AHA PREVENT risk models.

**Results:** The cohort was predominantly male (79.9%; mean age 44.6 ± 14.6 years), and abnormal ECG was the most common referral trigger (70.8%). Median 10-year risk was low by PREVENT and FRS and borderline elevated by PCE (PREVENT 2.7% [IQR 1.1-6.6], FRS 2.3% [IQR 0.5-7.1], and PCE 6.1% [IQR 2.4-9.2]). Among 72 participants with all scores, 51% were classified discordantly; PREVENT ASCVD agreed closely with FRS (κ=0.77) but poorly with PCE (κ=0.24-0.35), which consistently estimated higher risk. PCE was calculable in 52.6% versus 70.8% for PREVENT. Overall, 132 participants (85.7%) were cleared for deployment, while 8 (5.2%) were not physically qualified (NPQ); 6 of these NPQs were due to cardiovascular causes. Of 22 cardiac medical evacuations during the study period, 21 involved individuals without prior advanced cardiovascular screening. One screened individual was evacuated for symptoms subsequently found to have no cardiac cause.

**Conclusions:** Risk calculators disagree substantially in the fitness-selected population. We recommend PREVENT as the primary risk-stratification tool as it is the most contemporary derivation, incorporates renal function, is race-agnostic, extends to heart failure, applies from age 30, and agrees closely with FRS, easing transition. Targeted imaging, including coronary artery calcium scoring, may supplement intermediate-risk candidates.

## Introduction

The United States Antarctic Program (USAP) deploys thousands of personnel to Antarctica annually. The harsh environment and limited access to medical care present a major challenge when adverse medical events arise. Therefore, personnel require advanced medical screening prior to deployment to mitigate risk. Cold temperature exposure is a well-established physiologic stressor, triggering the sympathetic nervous system to release catecholamines and induce peripheral vasoconstriction, leading to elevated blood pressure and increased myocardial oxygen demand [1]. These hemodynamic changes promote a prothrombotic state that can precipitate myocardial ischemia, coronary vasospasm, and plaque instability [2,3]. Accordingly, the incidence of acute cardiovascular events increases significantly during colder months, a phenomenon documented in both hemispheres and more so in individuals who are not adapted to the cold [4,5]. The gravity of this risk is illustrated by a recent case report describing a 61-year-old male who suffered cardiac arrest due to an anterior ST-elevation myocardial infarction at McMurdo station, surviving with full neurologic recovery only after prolonged resuscitation and a 12-hour prehospital transport interval [6].

In recognition of these risks, the USAP, administered by the National Science Foundation (NSF) and supported medically by the Center for Polar Medical Operations (CPMO) at the University of Texas Medical Branch (UTMB) at Galveston, maintains a structured physical qualification (PQ) process. Standard pre-deployment screening includes a comprehensive history and physical exam and baseline laboratory studies. The 2025 Office of Polar Programs Physical Qualification guidelines [7] require a resting 12-lead ECG, repeated every 5 years for ages 40-49 years old, and annually for age 50 and over; stress testing is required only for winter-over personnel— every 2 years for ages 50-59 and annually for age 60 and over—or for any candidate with a Framingham Risk Score (FRS) >20%. Abnormal cardiovascular screening results are evaluated by a cardiologist. The first systematic description of the enhanced cardiovascular screening process was reported by Rodriguez-Lozano et al. in a retrospective analysis of 135 subjects evaluated between 2013 and 2017, in which 94.8% were ultimately cleared for deployment with no cardiac events reported among those who underwent advanced screening [8]. That study exclusively stratified participants’ risk based on their FRS and underscored the need for ongoing evaluation. Whether the current screening practices remain adequate in view of evolving cardiovascular risks has not been formally reassessed.

The FRS has been used historically and was derived from a community-based cohort but is known to have limited discriminatory value in low-risk populations [9]. The AHA/ACC Pooled Cohort Equations (PCE) expanded on FRS by incorporating race-specific variables which, like FRS, present limitations in this unique population [10,11]. Finally, the AHA PREVENT equations (2023) represent the most recent and comprehensive cardiovascular risk stratification tools by incorporating estimated glomerular filtration rate (eGFR), eliminating race as a variable, with improved calibration across a broader range of risk profiles compared to PCE and FRS [12]. Whether PREVENT’s theoretical advantages translate to USAP screening has not been evaluated.

The present study addresses these gaps through two aims. First, we characterize a contemporary cohort of 154 individuals referred for enhanced cardiovascular evaluation prior to USAP deployment, and compare the performance of the FRS, PCE, and PREVENT risk estimates and classification. Second, we evaluate the temporal trends in clearance rates, cardiovascular risk factors, and cardiac testing across three deployment cohorts (2018-2020, 2021-2023, and 2024-2026).

## Materials and Methods

### Study design and Cohort

We conducted a retrospective descriptive and exploratory analysis of individuals undergoing enhanced cardiovascular evaluation as part of pre-deployment medical screening for Antarctica deployment. The analytic dataset included participants evaluated between August 23, 2018, and April 2, 2026. Of 161 evaluation records, 4 records were excluded for incomplete data and 3 for duplicate entries, producing a final cohort of 154 unique individuals.

Cardiac medical evacuations occurring during the study period were identified from the CPMO medical evacuation records, in which each evacuation is categorized by CPMO clinical staff into a general medical category, including cardiac. The list of evacuations was cross-referenced by name against the analytic cohort to determine whether the evacuated individual had undergone prior advanced cardiovascular screening.

### Data sources and variables

The dataset included variables related to demographics, cardiovascular risk factors, past medical history, medications, vital signs, laboratory results, cardiovascular risk scores, ECG findings, stress testing, echocardiography, advanced cardiac testing, referral reason, deployment clearance status, outcomes, deployment exposure, and reviewer notes.

Outcome and deployment exposure-related variables were reviewed for feasibility. However, event outcomes, MEDEVAC status, event timing, deployment duration, deployment season, and activity level were largely unavailable or sparsely documented and were not used for primary analyses.

### Data cleaning and derived variables

Categorical variables were standardized, and numeric variables were parsed and screened. Medical evaluation date was used to derive evaluation year and evaluation period. Evaluation periods were grouped as 2018-2020, 2021-2023, and 2024-2026. Composite variables (current smoker; any CAD, arrhythmia, and major cardiac history; and echocardiography availability) were derived from their component fields.

### Cardiovascular risk scores

Analyses included multiple ten-year risk scores (PREVENT total CVD, ASCVD, CHD, Heart Failure, and Stroke; FRS; and the PCE). Because race is not recorded, the PCE, which requires race, was computed for every participant with both the White/other coefficient and the African American coefficient, and both estimates were carried through all analyses.

PREVENT, PCE (White or other), and PCE (African American) risk categories were defined as Low (<5%), Borderline (5% to <7.5%), Intermediate (7.5% to <20%), and High (≥20%). These thresholds are defined by the 2019 ACC/AHA guideline for 10-year ASCVD risk [13]. No analogous categorical thresholds have been established for PREVENT total CVD, CHD, heart failure, or stroke risk; the same cut points were applied to these outcomes by analogy for descriptive purposes. Framingham Risk Score categories were defined as Low (<10%), Intermediate (10% to ≤20%), and High (>20%). The Framingham Risk Score does not contain a borderline category.

### Statistical Analysis

Analyses were primarily descriptive. Continuous variables were summarized as mean ± standard deviation or median (IQR) and categorical variables were summarized as counts and percentages using available denominators. All analyses were complete-case; missing data were not imputed, and the denominator is reported for each variable so that missingness is explicit. Risk score availability, distribution, and category assignment were summarized among participants with calculable scores. ECG, stress testing, and echocardiography were performed selectively, so echo abnormalities were reported both overall and among those with available data.

Wilcoxon rank-sum tests were used for continuous variables, and Fisher’s exact tests were used for categorical variables. These comparisons were considered exploratory only. Agreement among risk models was assessed pairwise among participants with both scores available: Spearman rank correlation for continuous estimates and percent agreement with Cohen’s kappa for dichotomous low-versus elevated-risk classification (threshold <5% for PREVENT and PCE; <10% for FRS). Multivariable prediction models were not performed because of sparse outcome data, missing MEDEVAC data, and the small number of participants who were not cleared. All analyses were performed using R version 4.5.2.

## Results

### Cohort Characteristics

The final cohort comprised 154 unique participants evaluated between 2018 and 2026 (33 [21.4%] in 2018-2020, 63 [40.9%] in 2021-2023, and 58 [37.7%] in 2024-2026). Participants were mostly male (123, 79.9%), and middle-aged (mean 44.6 ± 14.6 years); baseline characteristics are summarized in Table 1. The most common risk factors were hyperlipidemia (24.3%), obesity (24.0%), and hypertension (14.3%), while diabetes (3.2%) and current smoking (4.5%) were uncommon. Any major cardiac history (coronary disease, arrhythmia, valvular disease, or syncope) was present in 11.0%, and any CAD history was rare (1.9%).

**Table 1.** Baseline Characteristics of the Study Cohort.

| Characteristic | Overall cohort<br>N = 154<sup>1</sup> |
| --- | --- |
| Age, years | 44.6 ± 14.6; N=154 |
| Sex |  |
| Female | 31/154 (20.1%) |
| Male | 123/154 (79.9%) |
| Height, cm | 175.8 ± 9.8; N=154 |
| Weight, kg | 84.0 ± 17.7; N=154 |
| Body mass index, kg/m<sup>2</sup> | 27.2 ± 4.9; N=154 |
| Heart rate, bpm | 73.2 ± 13.9; N=151 |
| Systolic blood pressure, mmHg | 123.1 ± 12.1; N=153 |
| Diastolic blood pressure, mmHg | 75.8 ± 8.4; N=153 |
| Hypertension | 22/154 (14.3%) |
| **Hyperlipidemia** | 37/152 (24.3%) |
| **Diabetes mellitus** | 5/154 (3.2%) |
| **Current smoker** | 7/154 (4.5%) |
| **Obesity history** | 37/154 (24.0%) |
| **Coronary artery disease** | 2/154 (1.3%) |
| **Myocardial infarction** | 2/153 (1.3%) |
| **Prior PCI** | 1/154 (0.6%) |
| **Prior CABG** | 0/154 (0.0%) |
| **Atrial fibrillation** | 3/154 (1.9%) |
| **Other arrhythmia** | 10/154 (6.5%) |
| **Heart failure** | 0/154 (0.0%) |
| **Valvular disease** | 3/152 (2.0%) |
| **Stroke/TIA** | 0/154 (0.0%) |
| **Chronic kidney disease** | 1/154 (0.6%) |
| **Syncope** | 4/154 (2.6%) |
| **Any CAD history** | 3/154 (1.9%) |
| **Any arrhythmia history** | 12/154 (7.8%) |
| **Any major cardiac history** | 17/154 (11.0%) |
| **Statin** | 14/154 (9.1%) |
| **ACE inhibitor/ARB** | 13/153 (8.5%) |
| **Beta blocker** | 8/153 (5.2%) |
| **Antiplatelet therapy** | 1/154 (0.6%) |
| **Anticoagulation** | 0/154 (0.0%) |
| **Antiarrhythmic therapy** | 1/154 (0.6%) |
| **Diabetes medication** | 5/154 (3.2%) |
| **Other lipid-lowering medication** | 2/153 (1.3%) |
| **Hemoglobin, g/dL** | 15.0 ± 1.3; N=139 |
| **Creatinine, mg/dL** | 0.9 ± 0.1; N=151 |
| **HbA1c, %** | 5.7 ± 0.9; N=9 |
| **Total cholesterol, mg/dL** | 188.6 ± 39.5; N=150 |
| **LDL cholesterol, mg/dL** | 112.0 ± 34.7; N=151 |
| **HDL cholesterol, mg/dL** | 56.5 ± 16.4; N=152 |
| **Triglycerides, mg/dL** | 104.0 ± 56.5; N=151 |
| **eGFR, mL/min/1.73 m<sup>2</sup>** | 96.1 ± 14.6; N=149 |
| **Glucose, mg/dL** | 92.8 ± 20.5; N=150 |

<sup>1</sup>Continuous variables are presented as mean ± SD with available N. Categorical variables are presented as n (%). Percentages are calculated using non-missing data.

### Cardiovascular risk scores

Risk score availability, distributions, and risk categories are summarized in Table 2. PREVENT and Framingham risk scores were calculable in most participants (70.8% and 74.7%), whereas the PCE was calculable in 52.6%, reflecting its narrower age range. Median 10-year risk was low by PREVENT and FRS and borderline by the PCE (median PREVENT total CVD 2.7%, PREVENT ASCVD 1.9%, Framingham Risk Score 2.3%, and PCE 6.1-7.2%), with most participants classified as low risk by PREVENT total CVD (65.1%) and FRS (87.0%) but only 32.5–43.2% by the PCE (Table 2). Risk scores generally increased with age across available models (Figure 1).

**Table 2.** Cardiovascular Risk Score Availability, Distribution, and Risk Categories.

| Risk score | Available<br>n/N (%) | Median<br>(IQR) | Low risk<br>n (%) | Borderline<br>n (%) | Intermediate<br>n (%) | High risk<br>n (%) |
| --- | --- | --- | --- | --- | --- | --- |
| PREVENT 10-year<br>total CVD risk | 109/154<br>(70.8%) | 2.7<br>(1.1–6.6) | 71<br>(65.1%) | 17<br>(15.6%) | 19 (17.4%) | 2 (1.8%) |
| PREVENT 10-year<br>ASCVD risk | 109/154<br>(70.8%) | 1.9<br>(0.9–4.0) | 93<br>(85.3%) | 10 (9.2%) | 6 (5.5%) | 0 (0.0%) |
| PREVENT 10-year<br>CHD risk | 109/154<br>(70.8%) | 1.0 (0.4–<br>2.2) | 104<br>(95.4%) | 4 (3.7%) | 1 (0.9%) | 0 (0.0%) |
| PREVENT 10-year<br>heart failure risk | 106/154<br>(68.8%) | 1.1 (0.4–<br>3.2) | 92<br>(86.8%) | 9 (8.5%) | 5 (4.7%) | 0 (0.0%) |
| PREVENT 10-year<br>stroke risk | 109/154<br>(70.8%) | 0.9 (0.4–<br>2.0) | 105<br>(96.3%) | 4 (3.7%) | 0 (0.0%) | 0 (0.0%) |
| Framingham Risk<br>Score | 115/154<br>(74.7%) | 2.3<br>(0.5–7.1) | 100<br>(87.0%) | 0 (0.0%) | 15 (13.0%) | 0 (0.0%) |
| PCE ASCVD risk<br>(White or Other) | 81/154<br>(52.6%) | 6.1<br>(2.4–9.2) | 35<br>(43.2%) | 13<br>(16.0%) | 30 (37.0%) | 3 (3.7%) |
| PCE ASCVD risk<br>(African American) | 80/154<br>(51.9%) | 7.2<br>(4.4–10.4) | 26<br>(32.5%) | 19<br>(23.8%) | 34 (42.5%) | 1 (1.2%) |

Risk category percentages are calculated among participants with available/calculable scores for each model. PREVENT, PCE ASCVD (White or Other), and PCE ASCVD (African American) prevention risk categories were defined as low <5%, borderline 5% to <7.5%, intermediate 7.5% to <20%, and high ≥20%; as no analogous categorical thresholds have been established for PREVENT CHD, heart failure, or stroke risk, these same thresholds were applied to those outcomes by analogy. Race was not available and PCE estimates were computed using both White/other and African American coefficients. Framingham Risk Score categories were defined as low <10%, intermediate 10% to 20%, and high >20%; borderline risk is not applicable for Framingham.

**Figure 1.**
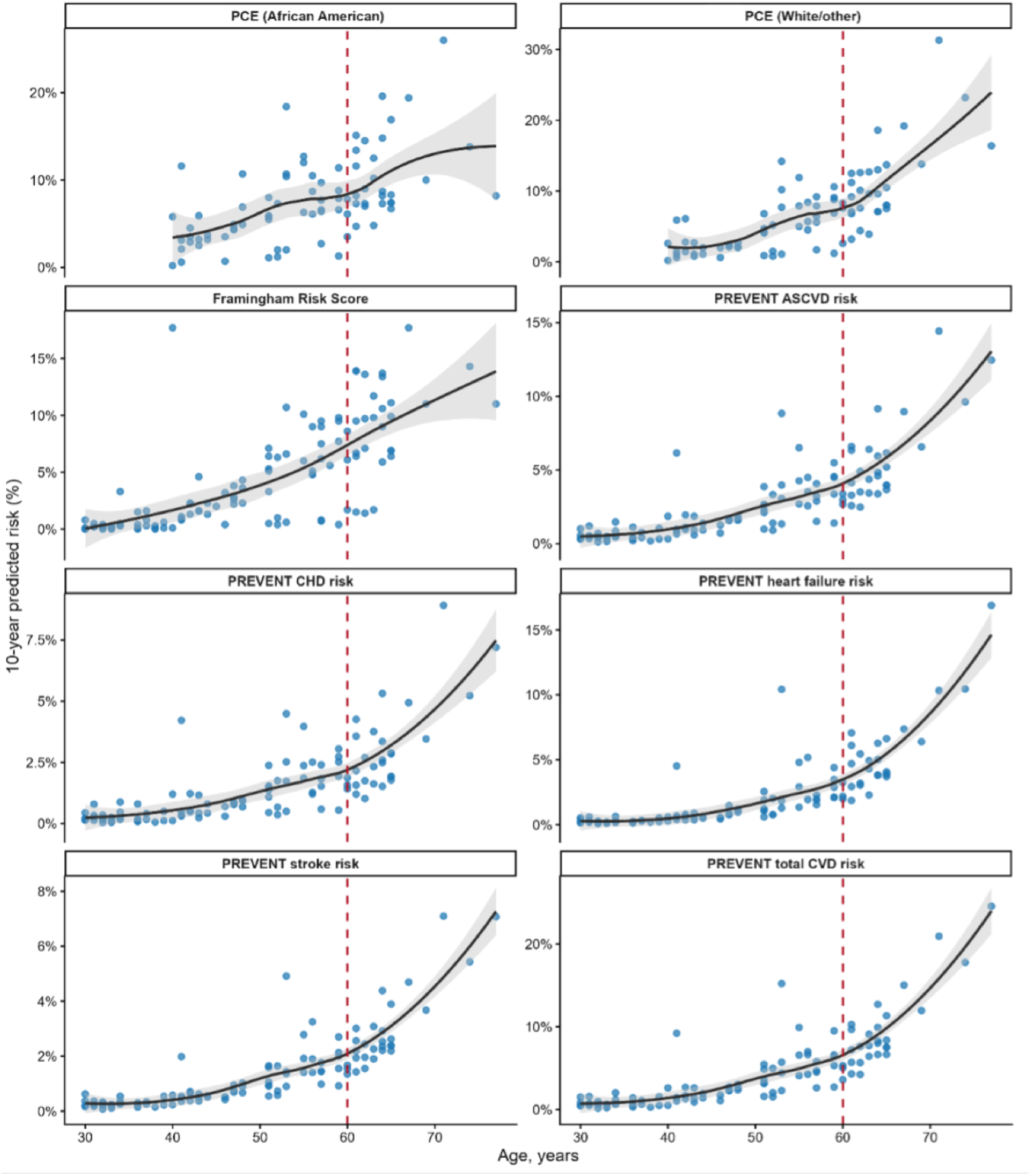
Cardiovascular Risk Scores by Age. Each panel shows individual participant-level 10-year predicted cardiovascular risk (%) plotted against age at evaluation for one risk model, with locally estimated scatterplot smoothing (LOESS) curve and shaded 95% confidence interval overlaid to illustrate the age-related trend. The dashed vertical line marks age 60 years, the threshold at which current USAP physical qualification guidelines require annual (rather than biennial) cardiac stress testing for winter-over personnel. Points represent participants with a calculable score for that model. Availability and distribution for each model are reported in Table 2.

### Reasons for referrals and cardiovascular testing

Referral reasons and ECG, stress testing, and echocardiographic findings are summarized in Table 3. Abnormal ECG was the dominant referral trigger (109/154, 70.8%); no other single reason exceeded 6% (the grouped “Other” category accounted for 10.4%). Stress testing was performed in 37 participants (24.0%), most commonly exercise ECG, with a median METs of 11.7 and symptoms during testing in 3 (8.1%; 1.9% overall). Echocardiography was available in 59 participants (38.3%); any abnormality was documented in 21 (13.6% overall, 35.6% of those imaged), most commonly valvular disease and left ventricular hypertrophy.

**Table 3.** Referral Reasons and Cardiac Testing Findings.

| Characteristic | n/N (%) |
| --- | --- |
| **Primary referral reason** |  |
| Abnormal ECG | 109 (70.8%) |
| Other | 16 (10.4%) |
| Abnormal stress test | 9 (5.8%) |
| Cardiology review | 8 (5.2%) |
| Bradycardia | 4 (2.6%) |
| ECG and Echo | 4 (2.6%) |
| Atrial Fibrillation | 2 (1.3%) |
| ECG and Cardiology review | 2 (1.3%) |
| **ECG findings** |  |
| Normal ECG | 13/154 (8.4%) |
| Not normal/abnormal ECG | 135/154 (87.7%) |
| ECG interpretation not documented | 6/154 (3.9%) |
| QTc, ms, median (IQR) | 413 (397–430); N=124 |
| **Stress testing** |  |
| Stress testing performed | 37/154 (24.0%) |
| No stress testing performed | 117/154 (76.0%) |
| Stress test METS, median (IQR) | 11.7 (10.3–13.4); N=34 |
| Symptoms during stress testing | 3/37 (8.1%) |
| **Stress test type among those tested** |  |
| ECG | 25 (67.6%) |
| Nuclear | 7 (18.9%) |
| Stress echo | 4 (10.8%) |
| Other/unclear | 1 (2.7%) |
| **Echocardiography** |  |
| Echo data available | 59/154 (38.3%) |
| LVEF, %, median (IQR) | 60 (55–60); N=48 |
| Any echo abnormality, overall cohort | 21/154 (13.6%) |
| Any echo abnormality, among echo-available | 21/59 (35.6%) |
| Left ventricular hypertrophy, among echo-available | 8/59 (13.6%) |
| Wall motion abnormality, among echo-available | 4/59 (6.8%) |
| Valve disease, among echo-available | 11/59 (18.6%) |
| Diastolic dysfunction, among echo-available | 4/59 (6.8%) |
| Percentages use non-missing denominators. Echocardiographic abnormalities are reported for the overall cohort and among participants with available echocardiographic data. Primary referral categories with a single participant each were grouped as “Other.” |  |

### Cardiovacular Screening Trends

Cardiovascular screening outcomes and temporal trends are summarized in Table 4. Overall, 132 participants (85.7%) were cleared, 8 (5.2%) were not cleared (6 for cardiovascular and 2 for non-cardiovascular reasons), and 14 (9.1%) were pending or unknown. Clearance declined across evaluation periods (97.0% to 88.9% to 75.9%). Over the same interval, obesity history and documented echocardiographic abnormality increased (obesity history 21.2% to 23.8% to 25.9%; echocardiographic abnormality 6.1% to 9.5% to 22.4%), though the latter is confounded by rising echocardiographic use.

**Table 4.**
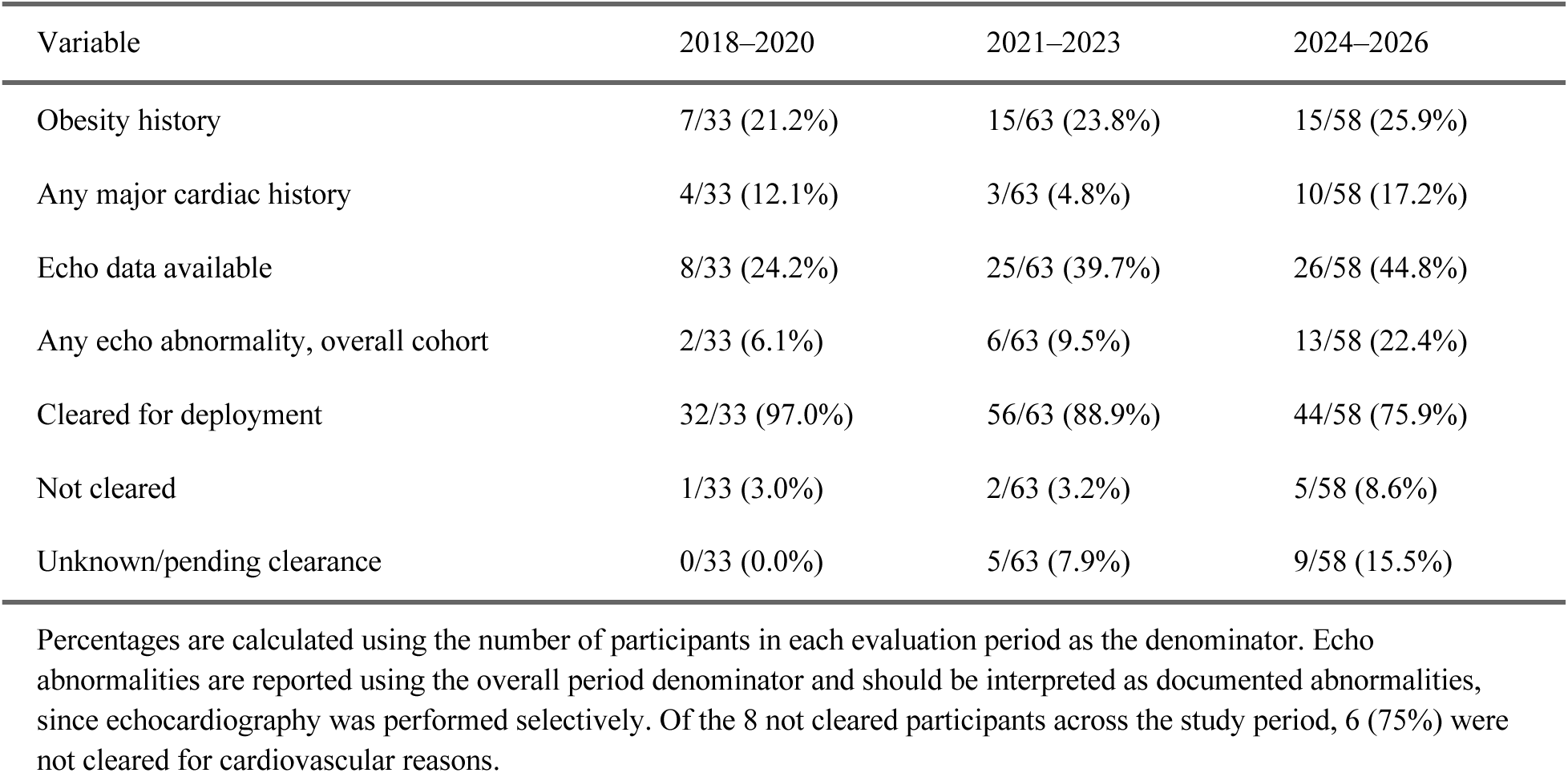
Risk Factor, Testing, Echocardiographic, and Clearance Trends Over Time.

During the study period, 22 cardiac medical evacuations occurred among deployed USAP personnel. Twenty-one involved individuals who had not undergone enhanced cardiovascular screening. The single evacuated participant who had been screened and cleared developed chest discomfort and dyspnea; evaluation in New Zealand identified no cardiac abnormality.

Denominator data for this analysis reflect the number of personnel physically qualified for deployment rather than confirmed deployment counts, as complete deployment figures were not available. Across the study period (2018-2026), 16,533 personnel were physically qualified for deployment. Using this figure, the 22 cardiac medical evacuations represent 0.13% of qualified personnel. The true rate among actually deployed personnel is necessarily slightly higher since a subset of physically qualified individuals do not deploy.

### Agreement and Disagreement Among Risk Calculators

The models ranked participants nearly identically (Spearman ρ 0.89-0.95 across pairwise comparisons) but agreed poorly on risk category (Table 5). Agreement on low versus elevated-risk classification was high between PREVENT and FRS (κ = 0.77) but only fair between PREVENT and PCE (κ = 0.24-0.35; 55-65% agreement). Discordance was essentially unidirectional: when models disagreed, PREVENT and FRS classified individuals as lower risk than the PCE in virtually every instance. Among the 72 participants with complete scores across all four models, only 35% were classified low risk by all four and 14% elevated by all four, leaving 51% discordant (Supplementary Table 5).

**Table 5.** Agreement and Reclassification Among Cardiovascular Risk Models.

| Model comparison | Paired, n | Spearman $\rho$ | Categorical agreement, %<sup>a</sup> | Cohen's $\kappa$ |
| --- | --- | --- | --- | --- |
| PREVENT ASCVD vs Framingham | 104 | 0.95 | 95 | 0.77 |
| PREVENT ASCVD vs PCE (White/Other) | 78 | 0.94 | 65 | 0.35 |
| PREVENT ASCVD vs PCE (African American) | 77 | 0.92 | 55 | 0.24 |
| Framingham vs PCE (White/Other) | 76 | 0.90 | 63 | 0.30 |
| Framingham vs PCE (African American) | 75 | 0.89 | 55 | 0.23 |
| PCE (White/Other) vs PCE (African American) | 80 | 0.91 | 88 | 0.74 |

<sup>a</sup> Categorical agreement and $\kappa$ reflect low- versus elevated-risk classification (threshold <5% for PREVENT and PCE models; <10% for Framingham). $\rho$ , Spearman rank correlation coefficient; $\kappa$ , Cohen's kappa.

### Exploratory comparisons

Exploratory comparison analyses were performed to compare participants based on stress test results, echocardiographic abnormalities, and clearance status (Supplementary Tables 1, 2, 3). These analyses were considered exploratory. The clearance comparison was limited by the small number of participants who were not cleared (N=8).

## Discussion

### Discordance Among Cardiovascular Risk Prediction Tools

The PREVENT and PCE models diverged most sharply (Table 2). For the same ASCVD endpoint, median risk was 1.9% by PREVENT versus 6.1-7.2% by the PCE, in addition to PCE placing 37-43% of the cohort in the intermediate risk category versus 5.5% by PREVENT ASCVD (17.4% by PREVENT total CVD). Agreement was high between the two PCE coefficient sets (κ = 0.74) but low between PREVENT/FRS and the PCE (κ = 0.23-0.35), indicating that model choice rather than race coefficient drives the observed discordance (Table 5). This pattern of PCE overestimation relative to PREVENT is consistent with recent validation studies attributing it to the PCE’s derivation from older, higher-event-rate cohorts [11]. The PREVENT equations were designed to address these shortcomings by excluding race as a predictor, incorporating eGFR, and validating against more diverse populations including lower-risk subgroups [12].

PCE overestimation, if used as the primary selection criterion for further cardiovascular testing or deployment restriction, would subject many low-risk candidates to unnecessary evaluation, cost, and logistical burden. This mismatch is large enough that current multi-society guidance defines materially different absolute thresholds across tools for the same nominal risk category, exemplified by “high risk” beginning at ≥20% by the PCE but ≥10% by PREVENT [14]. In contrast, the FRS model, which has been historically used in USAP screening guidelines, classified the largest proportion of the cohort as low risk compared to other models (Table 2). However, its known limitation in low-risk populations and the absence of variables such as eGFR limit its precision in this cohort [9]. This is particularly relevant given that renal function is closely associated with cardiovascular risk and may be vulnerable to extreme temperature exposure yet is entirely unaccounted for under the FRS or PCE [12,15]. Calculability also varied substantially (Table 2). PREVENT and FRS scores were obtainable in 70.8% and 74.7% of the cohort, respectively, whereas the standard PCE was calculable in only 52.6%. The PCE’s restricted age range and additional data requirement limit applicability here [13] and reinforce the advantages of PREVENT’s broader eligibility criteria and race-agnostic design.

In the echocardiographic subgroup analysis, both PREVENT and FRS scores were significantly higher among participants with any echocardiographic abnormality than among those without an abnormality. Median PREVENT risk was 4.7% versus 1.9% (p=0.021), and median FRS risk was 5.9% versus 1.1% (p=0.024) (Supplementary Table 2). Although neither group crossed conventional intermediate-risk thresholds, these associations suggest that both models capture a measurable gradient of underlying cardiovascular burden. However, they also demonstrate that calculated risk estimates cannot directly identify structural or functional abnormalities detectable only through imaging.

These findings support PREVENT as the preferred initial risk-stratification tool for USAP candidates, given its calculability, contemporary derivation, incorporation of renal function, and avoidance of race-based prediction. PREVENT should not, however, function as an independent determinant of deployment eligibility. Rather, it should complement the existing multimodal evaluation framework incorporating clinical history, electrocardiography, rhythm assessment, stress testing, and echocardiography when clinically indicated.

### Analogous Extreme Environment Populations

The consequences of cardiovascular risk misclassification are magnified by the remoteness of deployment. Paradoxically, efficient rescue from the South Pole can take longer than return from low-Earth orbit. In January 2026, NASA conducted its first medical early return from the International Space Station, returning Crew-11 safely back to Earth approximately one week after the event and roughly 10 hours after undocking [16]. Comparatively, evacuations during austral winter may be impossible for several months depending on conditions. Against these stakes, the discordance we report is not unique to polar medicine and recurs whenever traditional models are applied to occupational fit cohorts. A 2026 ACC/AHA scientific statement on the “tactical athlete” (military, fire service, and law enforcement) notes that no risk tool specific to this population exists and that available equations do not account for habitual fitness, which introduces imprecision for highly active individuals [14]. Additionally, there is a documented material difference in absolute thresholds across tools for the same nominal category [14]. Our cohort showed that approximately half of the subject pool would be reclassified based on the tool chosen but retained consistent ranking (Table 5; Supplementary Table 5). This is an expected outcome when models discriminate similarly but are calibrated differently, as even the best achieve a C-statistic of ∼0.80, leaving room for individual misclassification [17]. In an empirical comparison of five cardiovascular risk scores against 10-year observed outcomes in a multiethnic cohort, discrimination clustered narrowly (C-statistic 0.60-0.72) while calibration against real events varied as much as -21% to +154% depending on the model [18].

Other groups have responded to this challenge with supplemental imaging. NASA’s Astro-CHARM adds coronary artery calcium (CAC), hs-CRP, and family history for its astronauts [19]. A U.S. Navy program applied coronary CT angiography and CAC to older active-duty personnel and detected subclinical atherosclerosis in 54% despite a median 10-year ASCVD risk of 3.4%, prompting preventive therapy in about two thirds [20]. Aviation screening reveals a similar pattern to what we find here. Among asymptomatic pilots referred for cardiac evaluation, most had underlying cardiometabolic risk factors with excellent functional capacity showing mean METs of 11.9, and the most common echocardiographic abnormalities being valvular [21]. This profile is remarkably similar to the one we present (median METs of 11.7 and predominant valvular echocardiographic abnormalities). A review by the Royal Air Force revealed that aircrew referred for a cardiac abnormality who underwent cardiac MRI upgraded occupational status in 62%, returned 37% to unrestricted flying, and left only 8% undiagnosed [22]. Across environments and populations, the recurrent conclusion is that the discordance among calculators is expected for occupational cohorts. However, an equation derived from a broader and more contemporary population is the preferable primary tool if the output is treated as an initial stratification rather than a conclusion.

### Cardiovascular Screening Outcomes

Of 154 participants, 132 (85.7%) were cleared and 8 (5.2%) were not physically qualified, 6 due to cardiovascular causes (Table 4). The most distinct feature of NPQ participants was a higher prevalence of major cardiac history (37.5% vs 10.6%, p=0.045). The NPQ participants had lower median LDL than cleared participants (82.5 vs 112.0 mg/dL, p=0.040). This difference does not appear to reflect differential lipid-lowering therapy, as none of the NPQ participants were prescribed a statin and the NPQ group had no documented hyperlipidemia or CAD. The basis for this finding is unclear and, given the small NPQ group (n=8), should be interpreted as exploratory.

None of the three cardiovascular risk scores differed significantly between cleared and non-cleared participants. BMI, blood pressure, heart rate, and other conventional cardiovascular risk factors were also similar between the groups. These findings should not be interpreted as evidence that the risk equations lack clinical utility, because they were developed to estimate long-term cardiovascular event risk rather than occupational deployment eligibility. Instead, the results indicate that the clinical factors informing NPQ determinations were not fully represented by conventional risk estimates alone. The factors distinguishing disqualified participants in this cohort are captured by clinical history review, rhythm assessment, echocardiography, and other components of direct cardiovascular evaluation rather than by risk equations. This finding highlights the importance of a comprehensive multimodal evaluation rather than reliance on an isolated risk score or predetermined threshold.

### Screening Effectiveness and Evolving Risk Profile

Of the 22 cardiac medical evacuations during the study period, 21 involved individuals who had not undergone advanced cardiovascular screening. The one who had been cleared before deployment was evacuated after developing chest discomfort and dyspnea. Evaluation in New Zealand identified no cardiac abnormality. This observation is consistent with the USAP cohort from 2013 to 2017, in which no cardiac events were recorded among screened participants [8]. However, the absence of a formal comparison group, incomplete characterization of evacuated personnel, and the possibility of undisclosed preexisting conditions preclude causal conclusions. Accordingly, these findings support but do not definitively establish the effectiveness of enhanced cardiovascular screening.

The program also appears to be evaluating an increasingly complex referral population. The overall clearance rate of 85.7% was lower than the 94.8% reported in the prior series, and temporal analyses demonstrated a progressive decline in clearance (Table 4). Clearance rates decreased from 97.0% in the earliest cohort to 88.9% in the middle cohort and 75.9% in the most recent cohort. Over the same period, the proportion of participants with unknown or pending clearance status increased from 0% to 15.5%. This pattern may reflect increasing diagnostic complexity, changes in referral practices, incomplete follow-up, or the need for additional testing before a final clearance determination. Notably, obesity was not itself associated with lower clearance; participants with an obesity history were cleared at an equal or higher rate than those without in 2018–2020 and 2021–2023 and at a modestly lower rate in 2024–2026 (73.3% vs 76.7%) (Supplementary Table 4). The concurrent rise in obesity prevalence and decline in clearance therefore appear coincidental rather than causal, with the decline more likely explained by the rising proportion of pending determinations and expanded testing.

### Practical Recommendations

The AHA PREVENT equations should be adopted as the primary risk-stratification tool. They offer the most contemporary derivation, incorporate renal function, remain valid down to age 30, and extend prediction to heart failure, an outcome not captured by FRS or PCE (13.2% of this cohort was classified as borderline or intermediate risk by PREVENT heart failure). This transition carries low disruption, with ∼95% agreement between PREVENT and FRS on low versus elevated-risk classification (PREVENT vs FRS, Table 5). Although our data cannot directly evaluate imaging-based screening, analogous extreme-environment programs support its use as a logical extension. Extrapolating from NASA and U.S. Navy programs (Table 6), incorporating CAC scoring or coronary CT angiography for intermediate risk cases would address a documented weakness of risk-factor equations in fitness-selected populations [13,14,23,24]. This aligns with current multi-society guidance for occupationally fit populations, which recommends risk calculators incorporating CAC scoring for higher-risk task decisions while cautioning that CAC findings alone should not determine fitness-for-duty status and must be interpreted alongside functional capacity, symptoms, and ischemic evaluation [14]. This recommendation is not derived from outcomes measured here and would require prospective evaluation. The USAP screens thousands of candidates each year and even selective imaging would add radiation exposure, expense, and increased specialist utilization, so yield and cost-effectiveness in this specific population would need to be established before routine adoption. Further advanced imaging such as echo and MRI should remain reserved for those flagged by abnormal screening ECG or high calculated risk. The Royal Air Force shows that tiered imaging can resolve diagnostic uncertainty efficiently [22], and our data support a similar structure as participants referred for stress testing were reliably older (median age 56) and more comorbid. Those with echo abnormalities had significantly higher calculated risk, indicating the current abnormal ECG pathway is functioning as an appropriate filter.

**Table 6.**
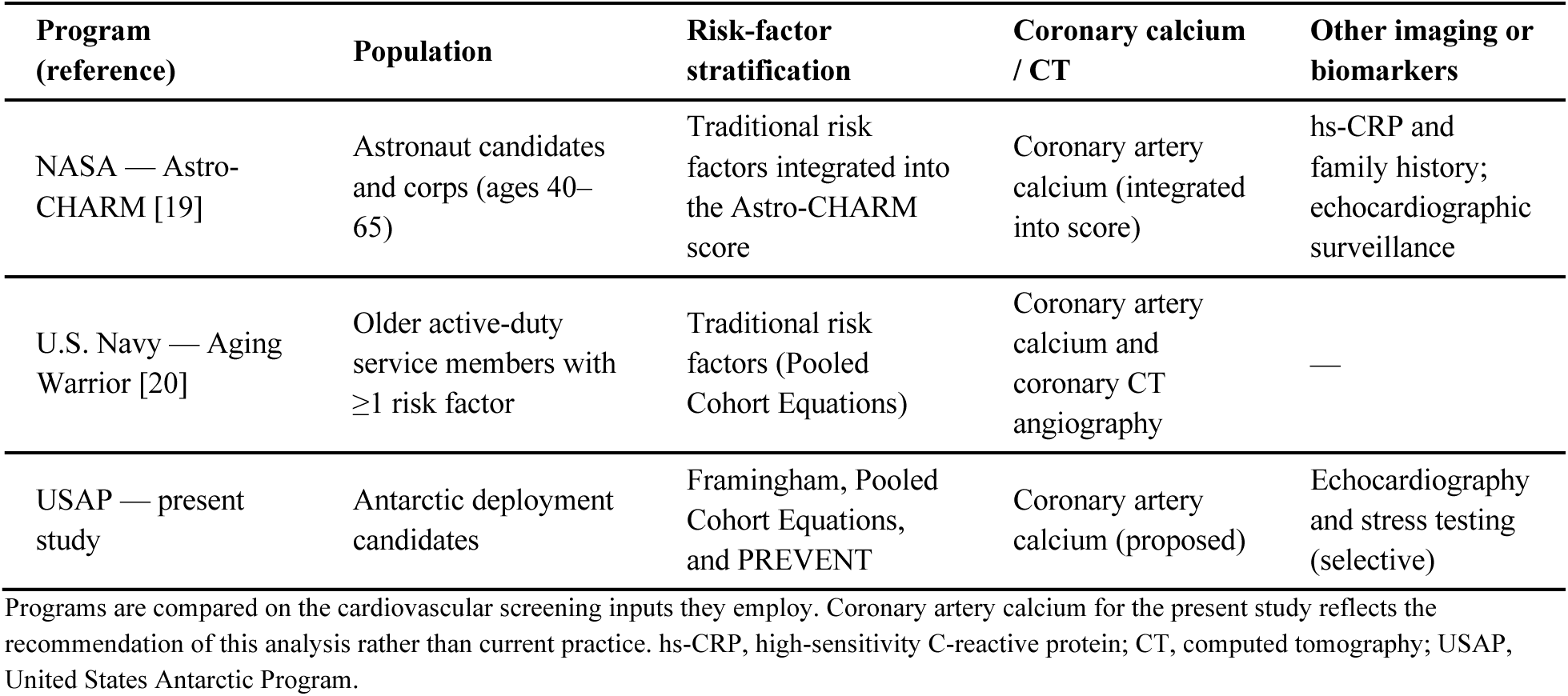
Cardiovascular Screening Across Analogous Extreme-Environment Occupational Programs.

### Limitations

This retrospective, single-program analysis examined a selected subset of candidates referred for enhanced cardiovascular evaluation, so the prevalence and risk distributions reported here should not be extrapolated to the general deployment population or to other programs. Cardiac medical evacuations were identified from the CPMO evacuation records and matched to the analytic cohort; however, detailed clinical review was not available for analysis so evacuation causality could only be characterized at the level of CPMO general classification. The single case involving a screened participant was individually reviewed. Our findings therefore describe discordance in classification, not differences in predictive accuracy. The high rank correlation but poor categorical agreement among tools points to differences in calibration and threshold placement rather than discrimination (Table 5). Model calculability and completeness were unequal: PREVENT, FRS, and PCE were calculable in 71%, 75%, and 53%, respectively. Race was not available as a variable, so PCE could not be applied as designed and was instead calculated with both coefficient sets. The not-cleared group was small (n=8), rendering all cleared versus not cleared comparisons exploratory and unsuitable for adjustment. Cardiac testing was performed selectively, and its use increased over time, so the apparent rise in echocardiographic abnormalities partly reflects greater ascertainment rather than a true increase in disease. Because history was self-reported by candidates with an incentive to be cleared, undisclosed symptoms or risk factors may have caused misclassification that no risk model or imaging protocol can detect. The denominator for cardiac medical evacuation rate reflects personnel physically qualified for deployment rather than personnel actually deployed, as the latter value was not available.

### Future Research

Future work should obtain confirmed annual deployment counts, rather than PQ counts, and aim to link risk scores and prior screening history to confirmed cardiac evacuations. This will aid in determining whether these models offer any prediction of actual events. Broader validation should follow through a prospective, multi-site design spanning other Antarctic programs or comparable occupations, which would also permit assessment of model performance by sex (women were only 20% of this cohort).

A hypothesis worth testing is whether ten-year event probability is the proper estimand for this setting. PREVENT, FRS, and PCE all estimate ten-year event probability under an assumption of prompt access to modern cardiac care, an assumption that does not hold during the austral winter, when evacuation may be delayed for weeks. What matters operationally is not whether a candidate will have an event within a decade, but whether that event will happen where rescue is measured in hours to weeks rather than minutes. A score built around that acute, access-limited framing rather than long-term prevention would better match the decision actually being made at clearance. Astro-CHARM and the Navy’s imaging-based program for older active-duty personnel suggest that occupation-specific tools are feasible once a population’s risk profile and resource constraints diverge enough from the general population [19,20].

Finally, non-disclosure will undermine any model regardless of calibration. Auditing how often self-reported history diverges from ECG, echocardiography, or laboratory findings could quantify the risk that currently escapes every tool examined here.

## Conclusion

USAP deployment candidates referred for enhanced cardiovascular evaluation were predominantly low risk by contemporary and traditional prediction models, but with significant discordance in absolute estimates, categorical classification, and calculability. PREVENT offers clear practical advantages; however, calculated risk scores did not distinguish cleared from non-cleared participants, whose outcomes were more closely associated with established cardiac history and abnormalities identified through direct cardiovascular evaluation. Only 1 of 22 cardiac evacuations involved an individual who had undergone advanced screening, and that case had no demonstrable cardiac cause, which is reassuring but should be interpreted cautiously given the study’s observational design. Collectively, these findings support the incorporation of PREVENT into USAP pre-deployment screening as an initial risk stratification tool while preserving individualized, multimodal cardiovascular evaluation for final clearance decisions.

## Statements and Declarations

### Author Contributions

**Daniel LeBlanc** – conceptualization, data curation, formal analysis, writing – original draft, writing – review and editing; **Abdul Qadeer** – conceptualization, data curation, formal analysis, writing – review and editing; **Shiva Raju Gollapally Krishna** – formal analysis, methodology, writing – original draft, writing – review and editing; **Andrew Forrester** – formal analysis, writing – original draft, writing – review and editing; **Anjni Patel** – data curation, writing – original draft; **Daniel Shi** – data curation, writing – original draft; **Eric Nagueh** – data curation, writing – original draft; **Jeremy Hsu** – data curation, writing – review and editing; **Neel Bhobe** – formal analysis, writing – original draft; **Diann Gaalema** – methodology, writing – review and editing, supervision; **James McKeith** – conceptualization, writing – review and editing, supervision; **Hani Jneid** – conceptualization, writing – review and editing, supervision; **Joseph Allencherril** – conceptualization, methodology, writing – review and editing, supervision, project administration. All authors reviewed and approved the final manuscript.

### Ethical Approval

This study was reviewed and approved by the University of Texas Medical Branch Institutional Review Board, protocol number 25-0347. It was conducted in accordance with the Declaration of Helsinki.

### Informed Consent

Given the retrospective design using existing clinical records, the requirement for informed consent was waived by the University of Texas Medical Branch Institutional Review Board.

### Generative AI Statement

During the preparation of this work the author(s) used Claude, developed by Anthropic, to assist in formatting of the tables and the references section. After using this tool/service, the author(s) reviewed and edited the content as needed and take(s) full responsibility for the content of the published article.

### Data Availability

The data that support the findings of this study are available from the corresponding author upon reasonable request. The data are not publicly available due to privacy restrictions on protected health information.

## Supplementary Tables

**Supplementary Table 1.**
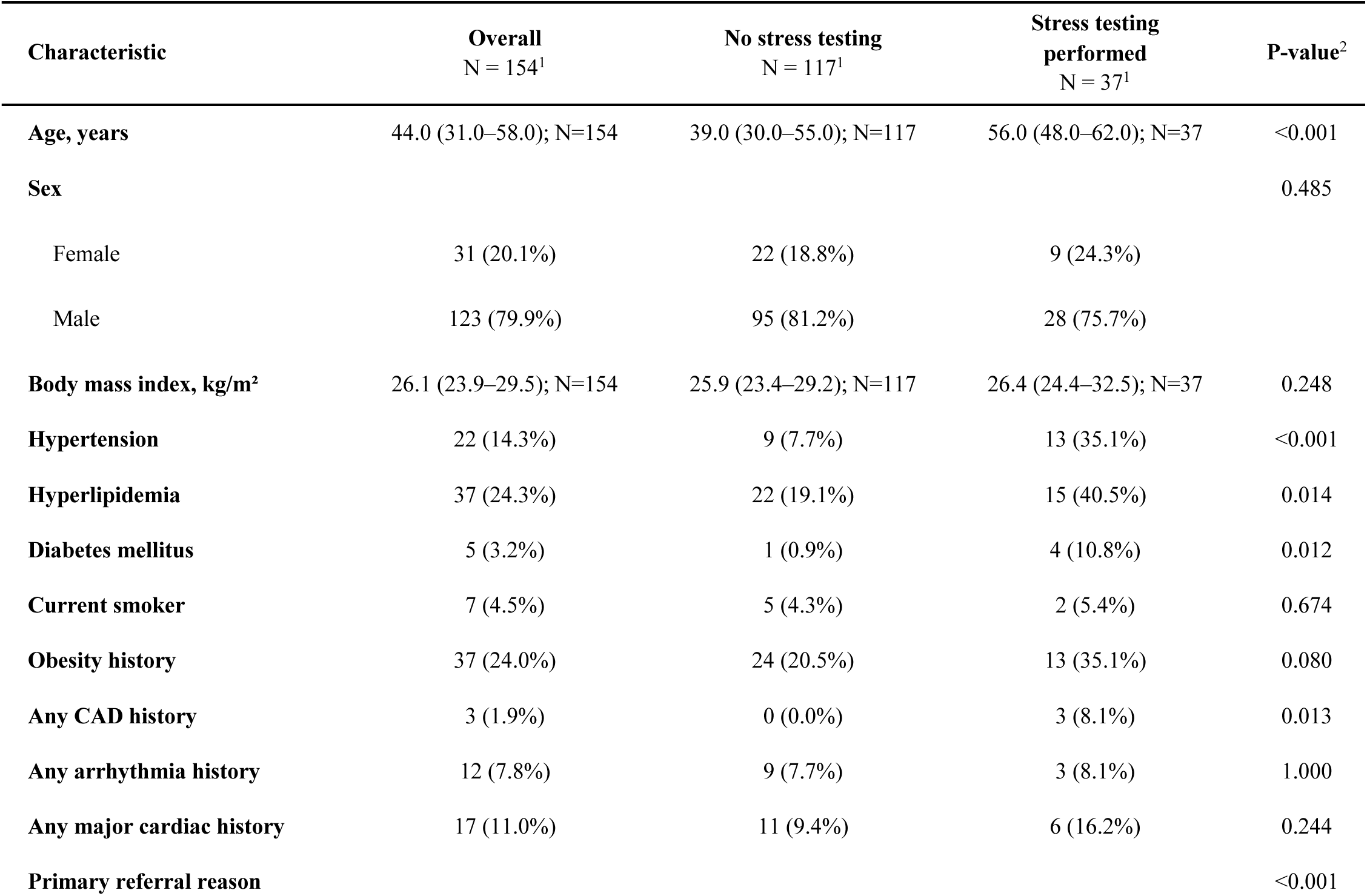
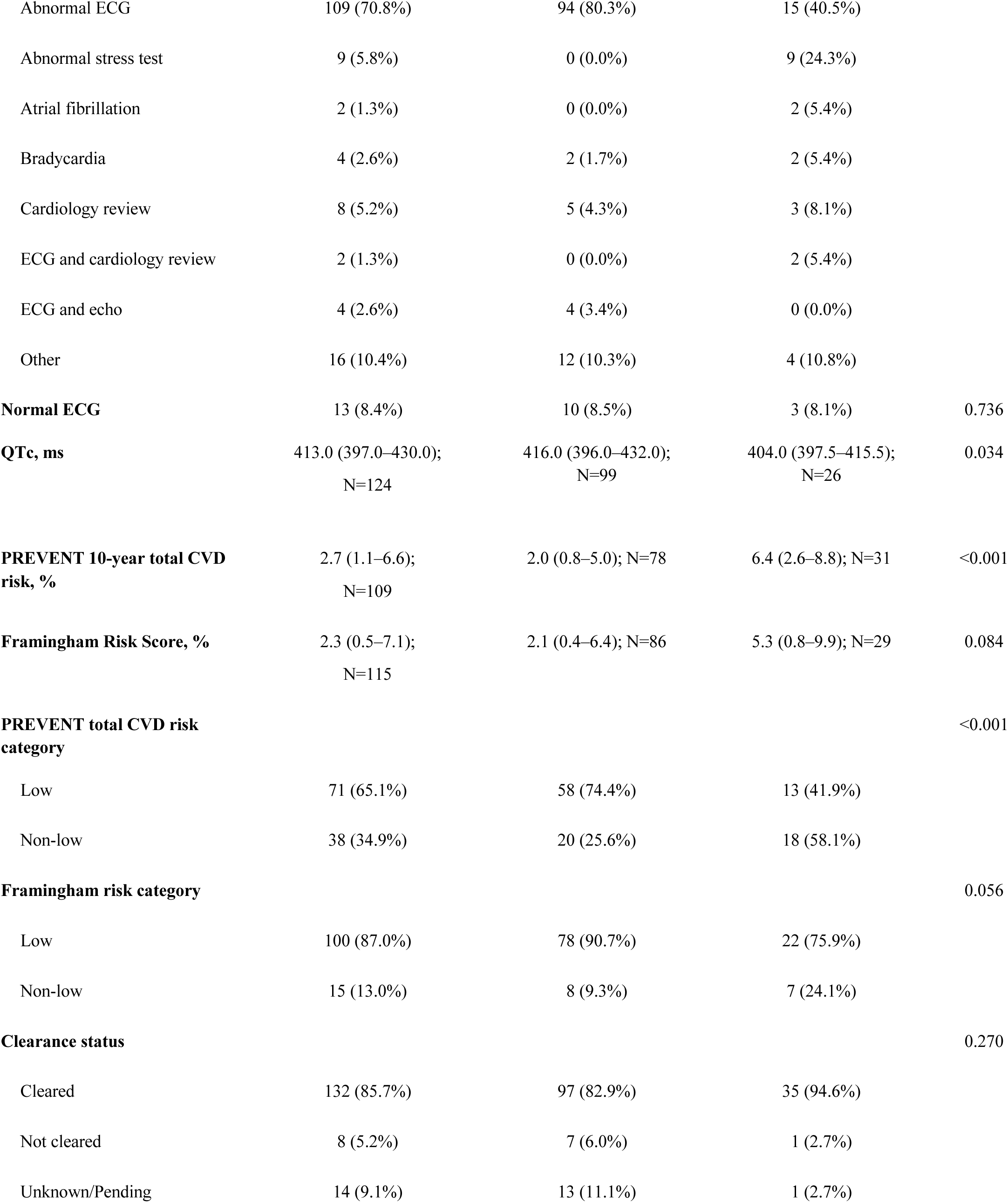
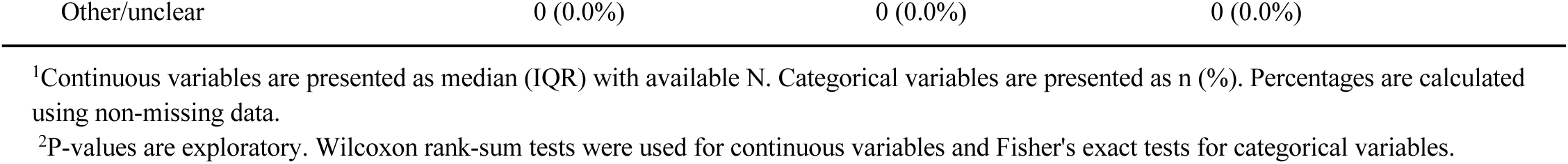
Exploratory Comparison of Participants by Stress Testing Status.

| Characteristic | Overall<br>N = 154<sup>1</sup> | No stress testing<br>N = 117<sup>1</sup> | Stress testing<br>performed<br>N = 37<sup>1</sup> | P-value<sup>2</sup> |
| --- | --- | --- | --- | --- |
| Age, years | 44.0 (31.0–58.0); N=154 | 39.0 (30.0–55.0); N=117 | 56.0 (48.0–62.0); N=37 | <0.001 |
| Sex |  |  |  | 0.485 |
| Female | 31 (20.1%) | 22 (18.8%) | 9 (24.3%) |  |
| Male | 123 (79.9%) | 95 (81.2%) | 28 (75.7%) |  |
| Body mass index, kg/m<sup>2</sup> | 26.1 (23.9–29.5); N=154 | 25.9 (23.4–29.2); N=117 | 26.4 (24.4–32.5); N=37 | 0.248 |
| Hypertension | 22 (14.3%) | 9 (7.7%) | 13 (35.1%) | <0.001 |
| Hyperlipidemia | 37 (24.3%) | 22 (19.1%) | 15 (40.5%) | 0.014 |
| Diabetes mellitus | 5 (3.2%) | 1 (0.9%) | 4 (10.8%) | 0.012 |
| Current smoker | 7 (4.5%) | 5 (4.3%) | 2 (5.4%) | 0.674 |
| Obesity history | 37 (24.0%) | 24 (20.5%) | 13 (35.1%) | 0.080 |
| Any CAD history | 3 (1.9%) | 0 (0.0%) | 3 (8.1%) | 0.013 |
| Any arrhythmia history | 12 (7.8%) | 9 (7.7%) | 3 (8.1%) | 1.000 |
| Any major cardiac history | 17 (11.0%) | 11 (9.4%) | 6 (16.2%) | 0.244 |
| Primary referral reason |  |  |  | <0.001 |
| Abnormal ECG | 109 (70.8%) | 94 (80.3%) | 15 (40.5%) |  |
| Abnormal stress test | 9 (5.8%) | 0 (0.0%) | 9 (24.3%) |  |
| Atrial fibrillation | 2 (1.3%) | 0 (0.0%) | 2 (5.4%) |  |
| Bradycardia | 4 (2.6%) | 2 (1.7%) | 2 (5.4%) |  |
| Cardiology review | 8 (5.2%) | 5 (4.3%) | 3 (8.1%) |  |
| ECG and cardiology review | 2 (1.3%) | 0 (0.0%) | 2 (5.4%) |  |
| ECG and echo | 4 (2.6%) | 4 (3.4%) | 0 (0.0%) |  |
| Other | 16 (10.4%) | 12 (10.3%) | 4 (10.8%) |  |
| **Normal ECG** | 13 (8.4%) | 10 (8.5%) | 3 (8.1%) | 0.736 |
| **QTc, ms** | 413.0 (397.0–430.0);<br>N=124 | 416.0 (396.0–432.0);<br>N=99 | 404.0 (397.5–415.5);<br>N=26 | 0.034 |
| **PREVENT 10-year total CVD risk, %** | 2.7 (1.1–6.6);<br>N=109 | 2.0 (0.8–5.0); N=78 | 6.4 (2.6–8.8); N=31 | <0.001 |
| **Framingham Risk Score, %** | 2.3 (0.5–7.1);<br>N=115 | 2.1 (0.4–6.4); N=86 | 5.3 (0.8–9.9); N=29 | 0.084 |
| **PREVENT total CVD risk category** |  |  |  | <0.001 |
| Low | 71 (65.1%) | 58 (74.4%) | 13 (41.9%) |  |
| Non-low | 38 (34.9%) | 20 (25.6%) | 18 (58.1%) |  |
| **Framingham risk category** |  |  |  | 0.056 |
| Low | 100 (87.0%) | 78 (90.7%) | 22 (75.9%) |  |
| Non-low | 15 (13.0%) | 8 (9.3%) | 7 (24.1%) |  |
| **Clearance status** |  |  |  | 0.270 |
| Cleared | 132 (85.7%) | 97 (82.9%) | 35 (94.6%) |  |
| Not cleared | 8 (5.2%) | 7 (6.0%) | 1 (2.7%) |  |
| Unknown/Pending | 14 (9.1%) | 13 (11.1%) | 1 (2.7%) |  |

<sup>1</sup>Continuous variables are presented as median (IQR) with available N. Categorical variables are presented as n (%). Percentages are calculated using non-missing data.
<sup>2</sup>P-values are exploratory. Wilcoxon rank-sum tests were used for continuous variables and Fisher's exact tests for categorical variables.

**Supplementary Table 2.** Exploratory Comparison by Echocardiographic Abnormality Among Participants With Available Echo Data.

| Characteristic | Overall<br>N = 59 | No echo<br>abnormality<br>N = 38 | Any echo<br>abnormality<br>N = 21 | P-value |
| --- | --- | --- | --- | --- |
| Age, years | 51.0 (33.5–59.5);<br>N=59 | 44.5 (31.2–56.8);<br>N=38 | 56.0 (40.0–62.0);<br>N=21 | 0.037 |
| **Sex** |  |  |  |  |
| Male | 47 (79.7%) | 29 (76.3%) | 18 (85.7%) |  |
| Female | 12 (20.3%) | 9 (23.7%) | 3 (14.3%) |  |
| Body mass index, kg/m<sup>2</sup> | 25.8 (23.1–29.1);<br>N=59 | 25.7 (23.4–28.6);<br>N=38 | 25.8 (23.0–31.1);<br>N=21 | 0.758 |
| Hypertension | 8/59 (13.6%) | 4/38 (10.5%) | 4/21 (19.0%) | 0.438 |
| Hyperlipidemia | 16/58 (27.6%) | 9/37 (24.3%) | 7/21 (33.3%) | 0.546 |
| Diabetes mellitus | 2/59 (3.4%) | 1/38 (2.6%) | 1/21 (4.8%) | 1.000 |
| Current smoker | 3/59 (5.1%) | 1/38 (2.6%) | 2/21 (9.5%) | 0.286 |
| Obesity history | 12/59 (20.3%) | 6/38 (15.8%) | 6/21 (28.6%) | 0.315 |
| Any CAD history | 2/59 (3.4%) | 1/38 (2.6%) | 1/21 (4.8%) | 1.000 |
| Any arrhythmia history | 4/59 (6.8%) | 3/38 (7.9%) | 1/21 (4.8%) | 1.000 |
| Any major cardiac history | 9/59 (15.3%) | 6/38 (15.8%) | 3/21 (14.3%) | 1.000 |
| **Primary referral reason** |  |  |  | 0.872 |
| Abnormal ECG | 39 (66.1%) | 25 (65.8%) | 14 (66.7%) |  |
| Abnormal stress test | 3 (5.1%) | 2 (5.3%) | 1 (4.8%) |  |
| Cardiology review | 7 (11.9%) | 4 (10.5%) | 3 (14.3%) |  |
| ECG and cardiology review | 2 (3.4%) | 2 (5.3%) | 0 (0.0%) |  |
| ECG and echo | 4 (6.8%) | 3 (7.9%) | 1 (4.8%) |  |
| Other | 4 (6.8%) | 2 (5.3%) | 2 (9.5%) |  |
| Normal ECG | 4/58 (6.9%) | 2/37 (5.4%) | 2/21 (9.5%) | 0.615 |
| QTc, ms | 408.5 (393.2–<br>423.8); N=54 | 408.5 (391.2–426.2);<br>N=34 | 409.5 (399.8–423.2);<br>N=20 | 0.585 |
| Stress testing performed | 17/59 (28.8%) | 11/38 (28.9%) | 6/21 (28.6%) | 1.000 |
| **Left ventricular ejection fraction, %** |  |  |  |  |
| 35 | 1 (2.0%) | 0 (0.0%) | 1 (5.0%) |  |
| 50 | 3 (6.0%) | 2 (6.7%) | 1 (5.0%) |  |
| 55 | 18 (36.0%) | 11 (36.7%) | 7 (35.0%) |  |
| 60 | 20 (40.0%) | 13 (43.3%) | 7 (35.0%) |  |
| 65 | 5 (10.0%) | 3 (10.0%) | 2 (10.0%) |  |
| 70 | 3 (6.0%) | 1 (3.3%) | 2 (10.0%) |  |
| Left ventricular hypertrophy | 8/59 (13.6%) | 0/38 (0.0%) | 8/21 (38.1%) | <0.001 |
| Wall motion abnormality | 4/59 (6.8%) | 0/38 (0.0%) | 4/21 (19.0%) | 0.014 |
| Valve disease | 11/59 (18.6%) | 0/38 (0.0%) | 11/21 (52.4%) | <0.001 |
| Diastolic dysfunction | 4/59 (6.8%) | 0/38 (0.0%) | 4/21 (19.0%) | 0.084 |
| PREVENT 10-year total CVD risk, % | 2.7 (1.1–6.6); N=47 | 1.9 (1.0–4.5); N=28 | 4.7 (2.5–7.6); N=19 | 0.021 |
| Framingham Risk Score, % | 2.3 (0.4–7.1); N=49 | 1.1 (0.4–4.8); N=30 | 5.9 (1.7–9.6); N=19 | 0.024 |
| **PREVENT total CVD risk category** |  |  |  | 0.109 |
| Low | 32 (68.1%) | 22 (78.6%) | 10 (52.6%) |  |
| Non-low | 15 (31.9%) | 6 (21.4%) | 9 (47.4%) |  |
| **Framingham risk category** |  |  |  | 0.285 |
| Low | 45 (91.8%) | 29 (96.7%) | 16 (84.2%) |  |
| Non-low | 4 (8.2%) | 1 (3.3%) | 3 (15.8%) |  |
| **Clearance status** |  |  |  | 0.492 |
| Cleared | 51 (86.4%) | 34 (89.5%) | 17 (81.0%) |  |
| Not cleared | 5 (8.5%) | 2 (5.3%) | 3 (14.3%) |  |
| Unknown/Pending | 3 (5.1%) | 2 (5.3%) | 1 (4.8%) |  |

<sup>1</sup>Analysis restricted to participants with available echocardiographic data. Continuous variables are presented as median (IQR) with available N. Categorical variables are presented as n (%).
<sup>2</sup>P-values are exploratory. Wilcoxon rank-sum tests were used for continuous variables and Fisher's exact tests for categorical variables

**Supplementary Table 3.**
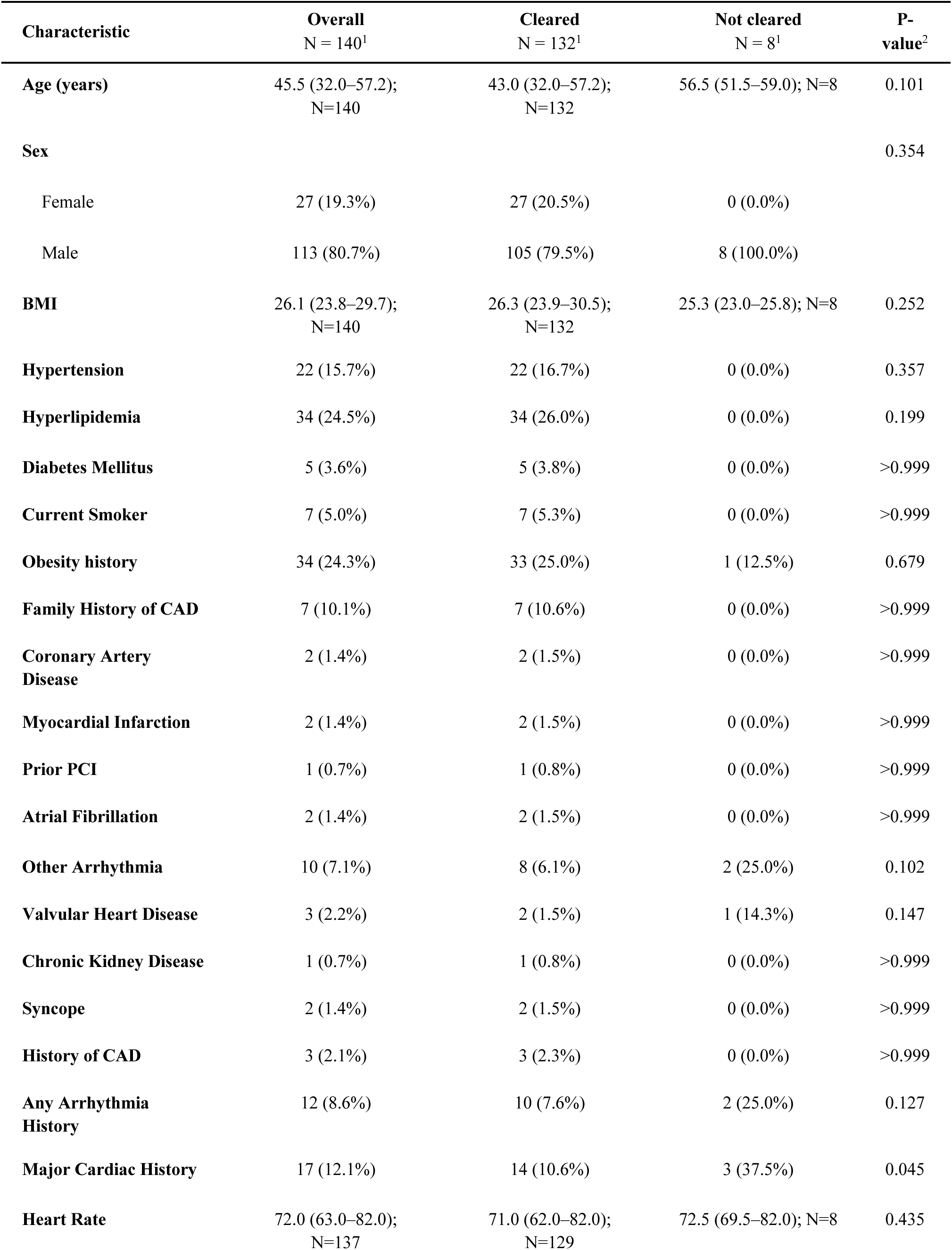
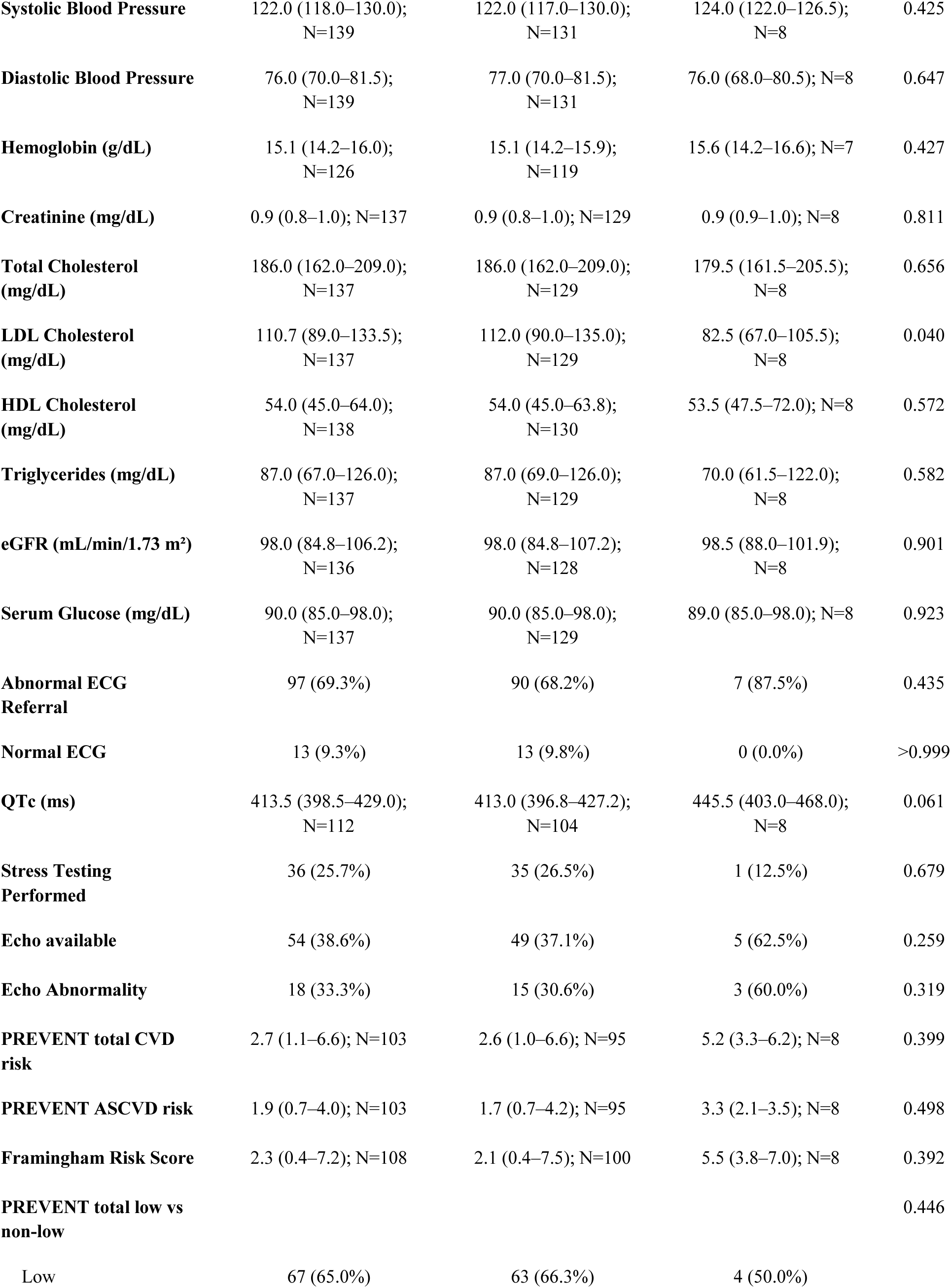
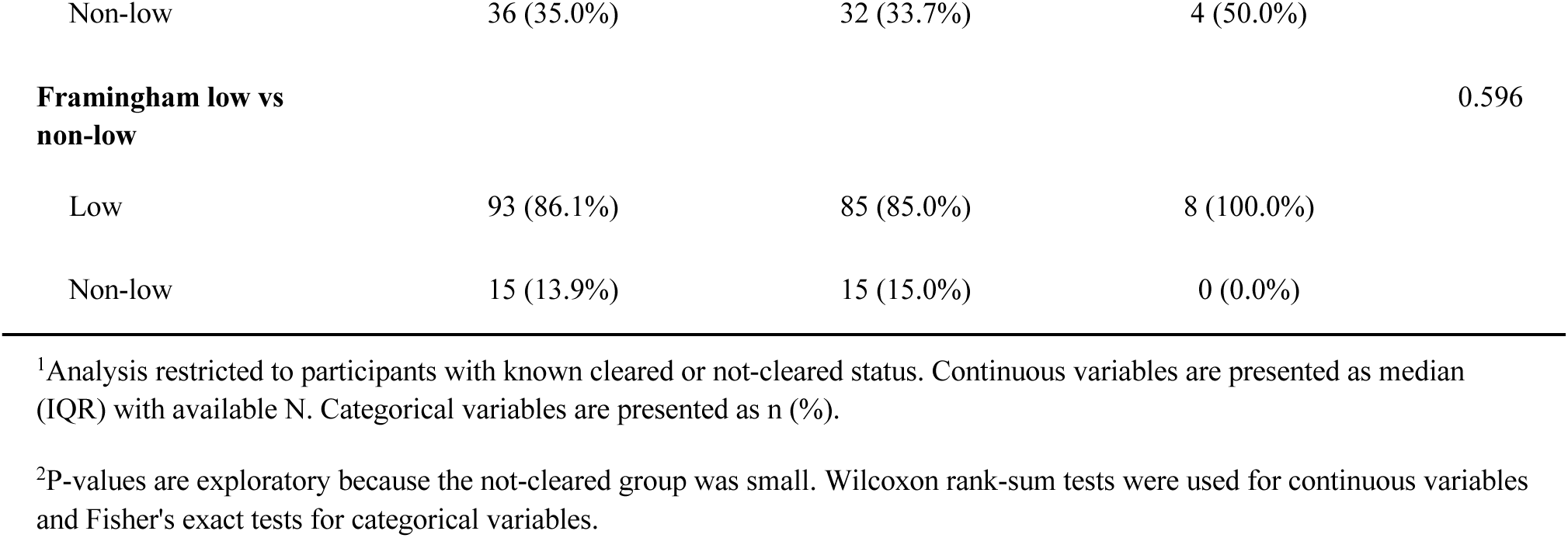
Exploratory Comparison of Participants by Clearance Status.

**Supplementary Table 4.** Clearance Status by Obesity History and Evaluation Period.

| Evaluation period | Obesity history | Total N | Cleared | Not cleared | Unknown/Pending |
| --- | --- | --- | --- | --- | --- |
| 2018-2020 | No obesity history | 26 | 25 (96.2%) | 1 (3.8%) | 0 (0%) |
| 2018-2020 | Obesity history | 7 | 7 (100%) | 0 (0%) | 0 (0%) |
| 2021-2023 | No obesity history | 48 | 41 (85.4%) | 2 (4.2%) | 5 (10.4%) |
| 2021-2023 | Obesity history | 15 | 15 (100%) | 0 (0%) | 0 (0%) |
| 2024-2026 | No obesity history | 43 | 33 (76.7%) | 4 (9.3%) | 6 (14.0%) |
| 2024-2026 | Obesity history | 15 | 11 (73.3%) | 1 (6.7%) | 3 (20%) |

Percentages are calculated within each obesity-history group and evaluation period.

**Supplementary Table 5.**
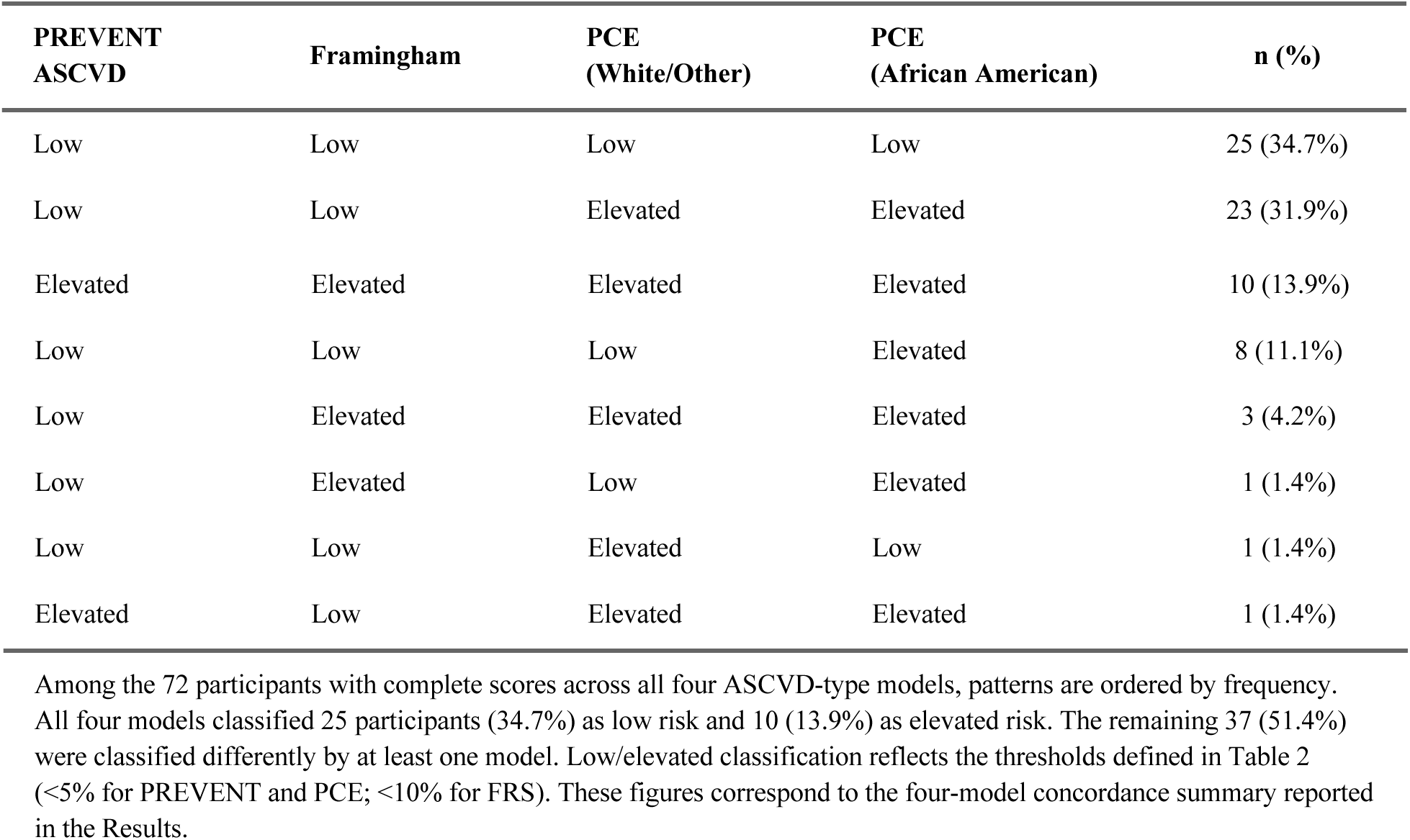
Cross-Tabulation of Risk Classification Across Four Cardiovascular Risk Models.

| PREVENT ASCVD | Framingham | PCE (White/Other) | PCE (African American) | n (%) |
| --- | --- | --- | --- | --- |
| Low | Low | Low | Low | 25 (34.7%) |
| Low | Low | Elevated | Elevated | 23 (31.9%) |
| Elevated | Elevated | Elevated | Elevated | 10 (13.9%) |
| Low | Low | Low | Elevated | 8 (11.1%) |
| Low | Elevated | Elevated | Elevated | 3 (4.2%) |
| Low | Elevated | Low | Elevated | 1 (1.4%) |
| Low | Low | Elevated | Low | 1 (1.4%) |
| Elevated | Low | Elevated | Elevated | 1 (1.4%) |

